# Chest pain presentations to hospital during the COVID-19 lockdown: lessons for public health media campaigns

**DOI:** 10.1101/2020.10.05.20203687

**Authors:** Amy V Ferry, Collette Keanie, Martin A Denvir, Nicholas L Mills, Fiona E Strachan

## Abstract

**Objective:** Emergency Department (ED) attendances with chest pain reduced during the COVID-19 lockdown. To understand factors influencing patients’ decisions to attend hospital, we performed a local service evaluation project in NHS Lothian.

**Methods:** We collated data on online searches and local clinical services on the number of ED presentations and chest pain clinic (CPC) referrals with suspected acute coronary syndrome between January and May 2020 and compared findings with the same period in 2019. We also carried out 28 semi-structured telephone interviews with patients who presented with chest pain during lockdown and in patients with known coronary heart disease under the outpatient care of a cardiologist in April and May 2020. Interviews were audio recorded and salient themes and issues documented as verbatim extracts.

**Results:** Online searches for the term “chest pain” doubled after 01/03/2020, peaking in week commencing 22/03/2020 and returning to 2019 levels during April 2020. In contrast, chest pain presentations to ED and CPC decreased, with the greatest reduction in the final week of March 2020 (128 v 287 (average weekly ED attendance 2019), and 6 v 23 (average weekly CPC referral 2019)). This aligned with key government messages to ‘Protect the NHS’ and the ‘NHS is open’ campaign. Patient interviews revealed three main themes; 1) pandemic help-seeking behaviour2) COVID-19 exposure concerns; 3) favourable Hospital experience if admitted.

**Conclusions:** Dynamic monitoring of public health and media messaging should evaluate public response to healthcare campaigns to ensure the net impact on health, pandemic and non-pandemic related, is optimised.

**What is already known about the subject?:** Reports from around the world revealed a decrease in the numbers of patients attending hospital for serious health complaints such as chest pain during the lockdown restrictions imposed by governments to decrease the spread of SARS-CoV-2.

**What does this study add?:** This service evaluation project has provided insight into how patients experiencing chest pain made the decision to attend hospital for assessment during this period. It has revealed how the pandemic shaped help-seeking practices, how patients interpreted their personal vulnerability to the virus, and describes patient experience of attending hospital for assessment during this time.

**How might this impact on clinical practice?:** Careful monitoring of the public response to health care messaging campaigns should be a key part of a pandemic strategy and careful adjustment of messaging, in a dynamically responsive way, should be considered in future.

## Introduction

Symptoms suggestive of acute coronary syndrome are one of the most common reasons for Emergency Department presentation [1]. Reports from around the world have described a considerable decrease in the numbers of patients presenting to the Emergency Department with chest pain coinciding with the arrival of COVID-19 and associated lockdown restrictions [2]. Scotland is no different with NHS performance indicators reporting that Emergency Departments experienced a substantial reduction in attendances during government advice recommending strict social distancing during the COVID-19 pandemic [3]. This has led to concerns that patients with significant illness such as myocardial infarction may not be attending hospital. National data also reveal excess total mortality that is not completely attributable to COVID-19 [4] suggesting additional public harm may be resulting from decreasing Emergency Department attendances for non-COVID associated illness.

Scotland entered lockdown on 23^rd^ March 2020 with advice to ‘stay home, protect our NHS, save lives’. Patients admitted to acute medical units in NHS Lothian, Scotland, during the first 31 days after lockdown were of higher medical acuity and had a higher risk of inpatient mortality when compared to patients in the same period in the preceding 5 years [5]. This suggests that patients may not have been seeking and accessing healthcare in the same way as prior to the COVID-19 outbreak. It is imperative that patients with chest pain seek professional healthcare assessment for what could potentially be a medical emergency which, if not treated, can lead to life threatening complications such as heart failure, arrhythmia and death.

We aimed to explore how the COVID-19 pandemic and public health advice affected chest pain presentations and help-seeking behaviour in NHS Lothian.

## Methods

We collected data on online searches for the term ‘chest pain’ from 01 January 2020 to 31 May 2020 using Google Trends with data presented as a percentage change from previous month. In addition, we collected local service data on the number of patients attending the Emergency Departments (ED) in NHS Lothian with suspected acute coronary syndrome as well as the number of GP referrals to the Chest Pain Clinic at the Royal Infirmary of Edinburgh where patients can access an expedited outpatient assessment for new onset chest pain with worsening angina symptoms. These data were compared to the same time period from the previous year. Key dates used in the analysis include: March 2^nd^, first confirmed COVID-19 case in Scotland; March 23^rd^, lockdown restrictions began; April 4^th^ media reports of reduced presentations to hospital; April 26^th^, public health campaign to attend hospital for chest pain.

Semi-structured telephone interviews were also conducted with patients attending hospital for the assessment of suspected acute coronary syndrome between the 17 April 2020 and 08 May 2020 (n=21). Participants were identified using a previously described order request for cardiac troponin used in all patients with suspected acute coronary syndrome and by review of the electronic patient record [6]. A further 7 telephone interviews were conducted with patients with known coronary heart disease under the care of a cardiologist as an outpatient. The rationale for including these patients was to try to capture the experience of patients who may have experienced chest pain but may not have chosen to attend hospital during this period. A topic guide (appendix 1) was developed by the study team comprising of two consultant cardiologists and two nurses. All interviews were conducted by AVF (a female cardiology research nurse with experience in qualitative interviewing) and lasted 5 to 37 minutes (mean 12 minutes). Recruitment continued until saturation was achieved and additional interviews did not yield new insights [7]. Participants were asked to talk about how they made the decision to attend hospital for assessment and whether coronavirus had impacted that decision making process. Participants from the outpatient clinic lists were asked if they had experienced symptoms in the preceding two months and whether coronavirus had impacted how they dealt with those symptoms. All those contacted were happy to participate and for the conversation to be recorded. An interpretivist approach was taken to data analysis. Detailed notes were written with salient issues noted whilst listening to the whole voice files. Notes were read repeatedly searching for patterns of meaning in the data. Verbatim extracts were documented as necessary to illustrate themes arising [8]. Emerging themes were discussed with FS and MAD. This project was conducted as a service evaluation project and registered with the local cardiology quality improvement team according to local practice in NHS Lothian. It was reviewed by South East Scotland Research Ethics Service where it was assessed as not requiring further ethical review.

### Patient and public involvement

The project was conceptualised through discussion with patients admitted to a cardiology ward during the COVID-19 pandemic. Key ideas for interview questions were developed by consultation with the patient group.

## Results

### Online symptom searches

Google trends searches in Scotland for the term “chest pain” doubled from March 1^st^, reaching a peak between March 22^nd^ and 29^th^ (101% change from previous months average) and returned to baseline levels over the next 4 weeks. Data from 2019 revealed a consistent use of the search term “chest pain” at 46%-68% of the peak activity seen in 2020 over the time matched period (**Figure 1**).

**Figure 1.**
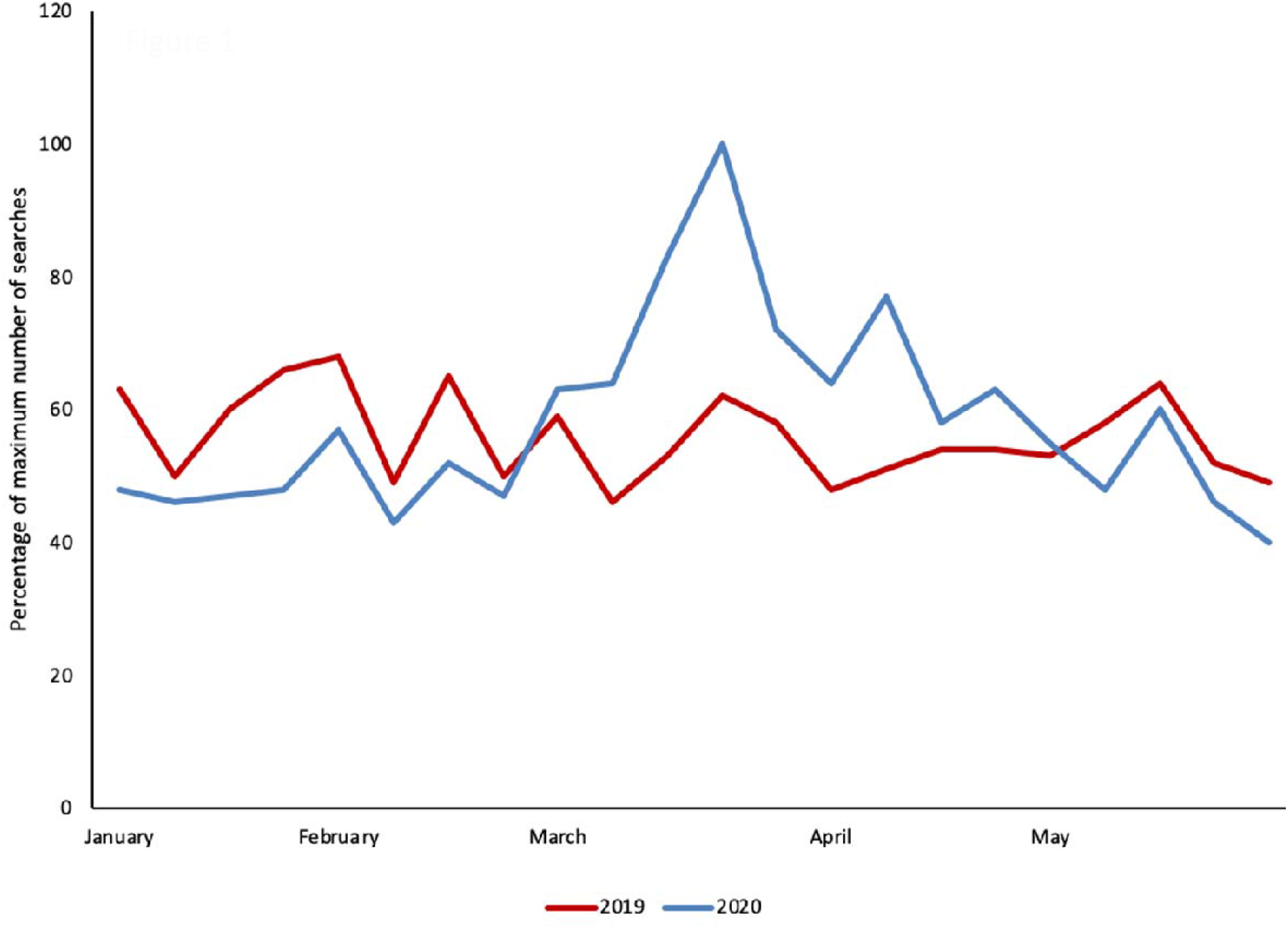
Online searches for ‘chest pain’ in Scotland. Chart showing number of searches for the term ‘chest pain’ expressed as percentage of maximum searches from January to May 2019 and 2020 in Scotland

### Emergency Department attendance and referrals to chest pain clinic

The average weekly number of ED attendances between January and May with suspected acute coronary syndrome was 287 in 2019 dropping to 233 in 2020. The lowest number of attendances per week (128) was seen in the last week of March 2020 (**Figure 2**). We observed a similar decrease in referrals to the CPC with an average of 23 per week in 2019 dropping to 16 per week in 2020. The lowest number was 6 referrals in the last week of March and referral rate remained between 6 and 11 per week throughout April 2020.

**Figure 2.**
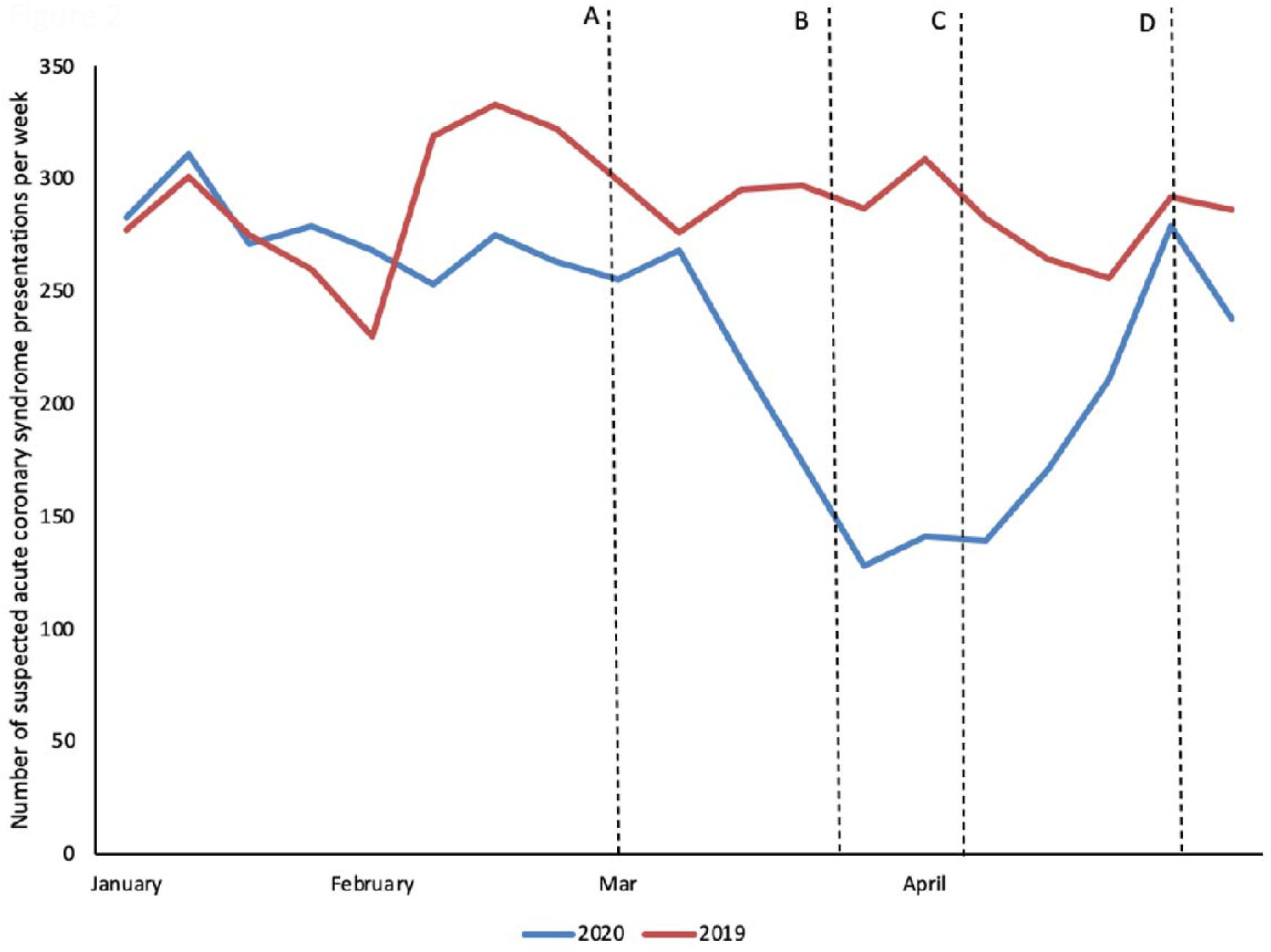
Emergency Department presentations to NHS Lothian per week with suspected acute coronary syndrome from January to May 2019 and 2020. Key dates are denoted by ‘A’ first confirmed COVID-19 case in Scotland 02 March 2020, ‘B’ lockdown enforcement 23 March 2020, ‘C’ British Heart Foundation and media reports of decreased hospital presentation 04 April 2020, ‘D’ ‘The NHS is open’ campaign launched 26 April 2020

### Characteristics of interview participants

Interview participants were aged between 39 and 88 years and 54% were female. 14 participants had an admission troponin greater than the diagnostic threshold for myocardial infarction, 7 had an admission troponin less than the diagnostic threshold for myocardial infarction and 7 were under the care of a cardiologist as an outpatient.

### Semi-structured interview results

Telephone conversations revealed three main themes; pandemic shaping of help seeking behaviour, COVID exposure concern, and hospital experience. These are summarised in **Figure 3** with possible implications for practice.

**Figure 3.**
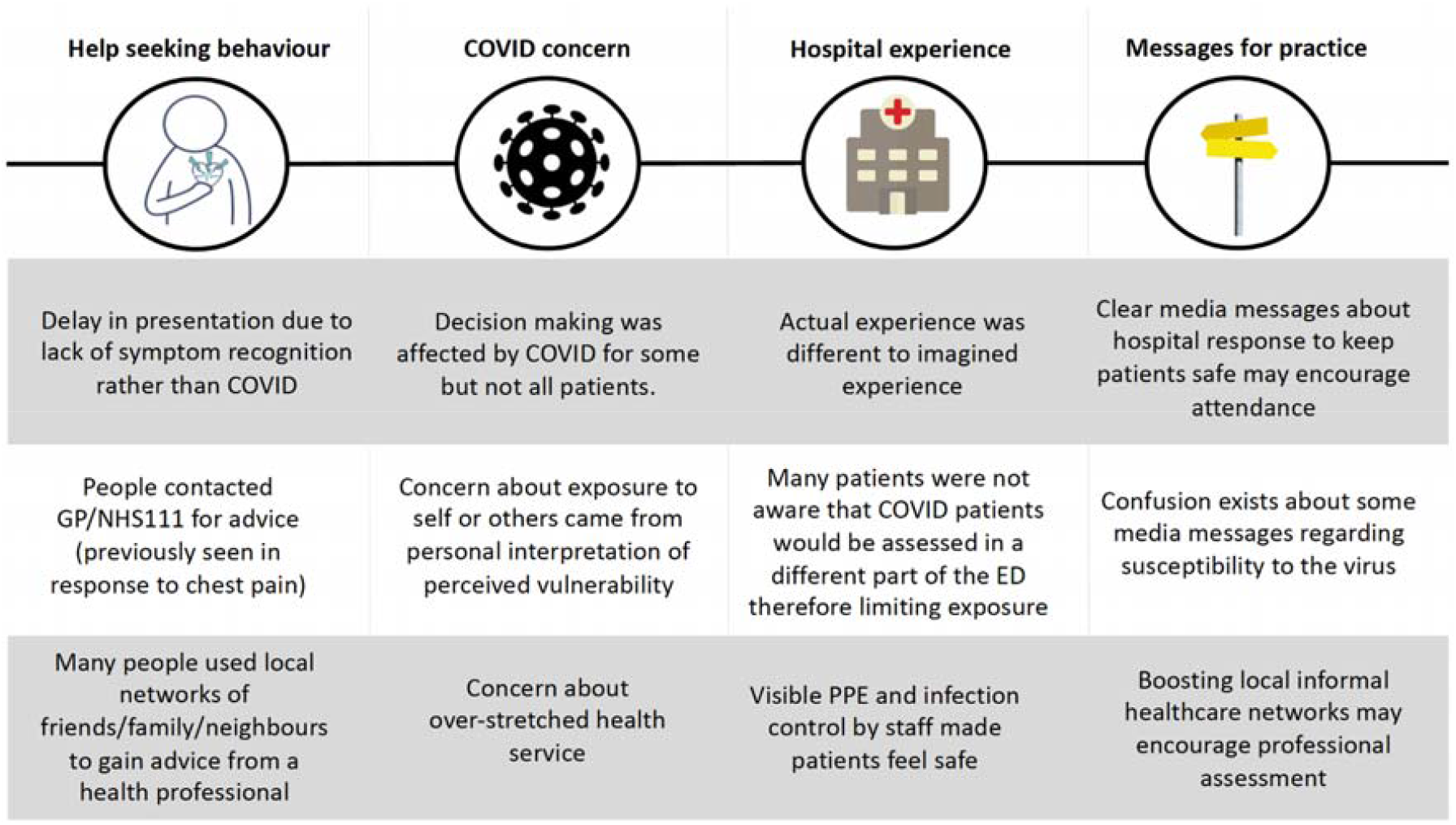
Summary of main interview themes and lessons for practice.

### Pandemic shaping of help seeking behaviour

Participants revealed a staged response to seeking help. Firstly, they reported performing a symptom appraisal which could lead to symptoms being attributed to other causes, for example indigestion, due to the transient nature of their symptoms or lack of severity. Persistence of symptoms triggered discussion with local neighbourhood networks and family to decide on the next course of action. It was noted that for the participants in this study, these networks often included a healthcare professional. The outcome of this discussion commonly involved phoning a GP or the NHS out of hours service (NHS24) for preliminary assessment (13/20 participants). No barriers to accessing these services were reported. Only 3/20 participants telephoned the Scottish Ambulance Service directly without additional assessment, with the remainder self-presenting to the Emergency Department.

Some participants reported that gaining access to a physical examination was difficult. Consultations tended to be telephone based which were sometimes viewed as inadequate due to difficulty in describing symptoms over the telephone to a doctor with whom they did not have a relationship. Other participants did have access to face-to-face primary care appointments but similarly these were not viewed positively due to lack of physical examination.

*“The only thing he did was put a thing on my finger and take my temperature”* (participant 20)

One patient described accessing three GP phone consultations and an out of hours appointment where she had blood pressure and oxygen saturations recorded but no ECG despite describing chest, arm and jaw pain. This patient subsequently self-presented to the Emergency Department and was diagnosed with acute myocardial infarction. This participant illustrates her experience by saying; *“They were paranoid about COVID*.*”* (participant 8)

### COVID-19 exposure concern

Participants were asked how coronavirus had affected their decision making of whether to attend hospital. Participants differed in their response with some stating their decision making was completely unaffected. Interpretations of this theme are illustrated using participant quotes in **Table 1**.

**Table 1.**
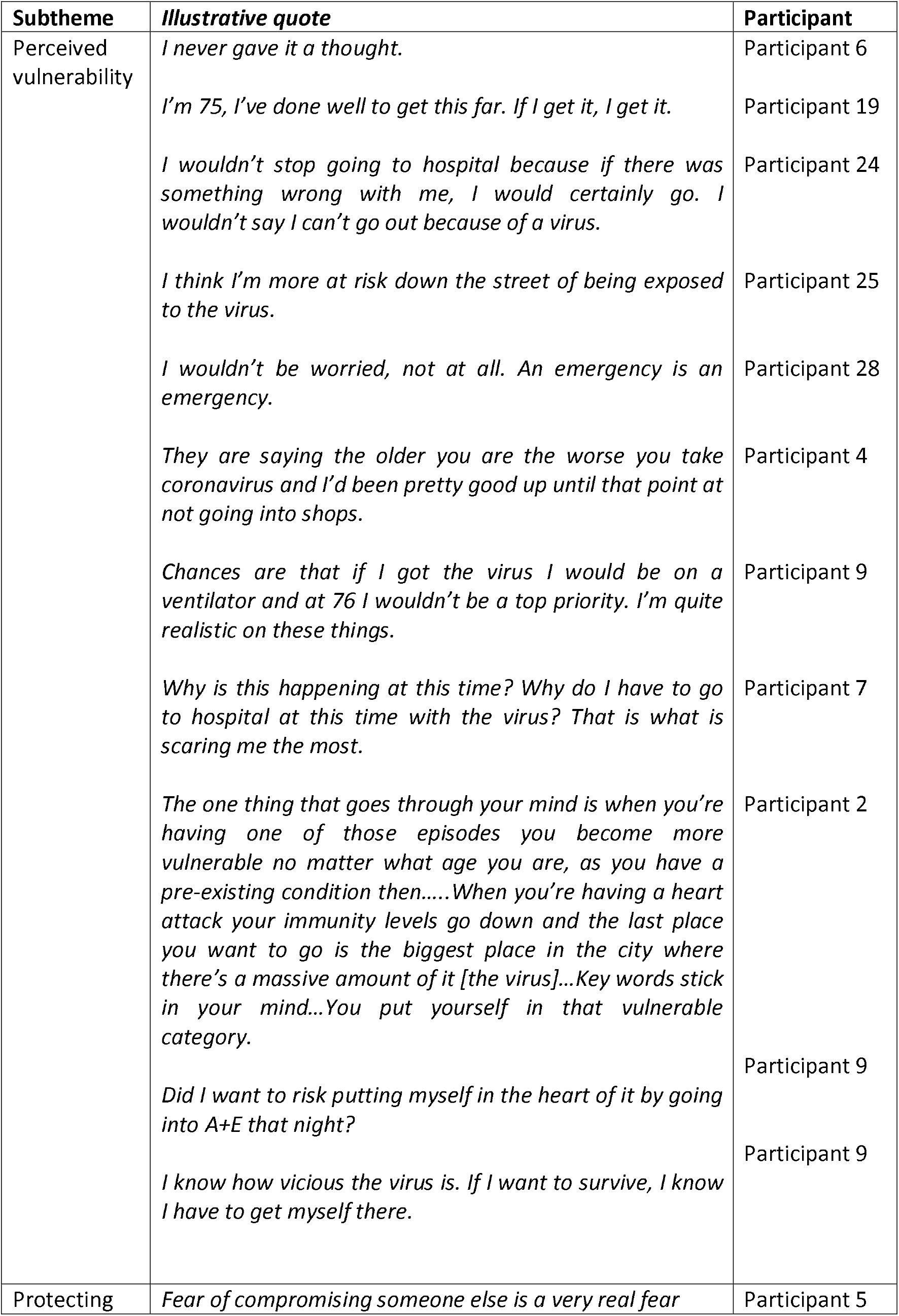
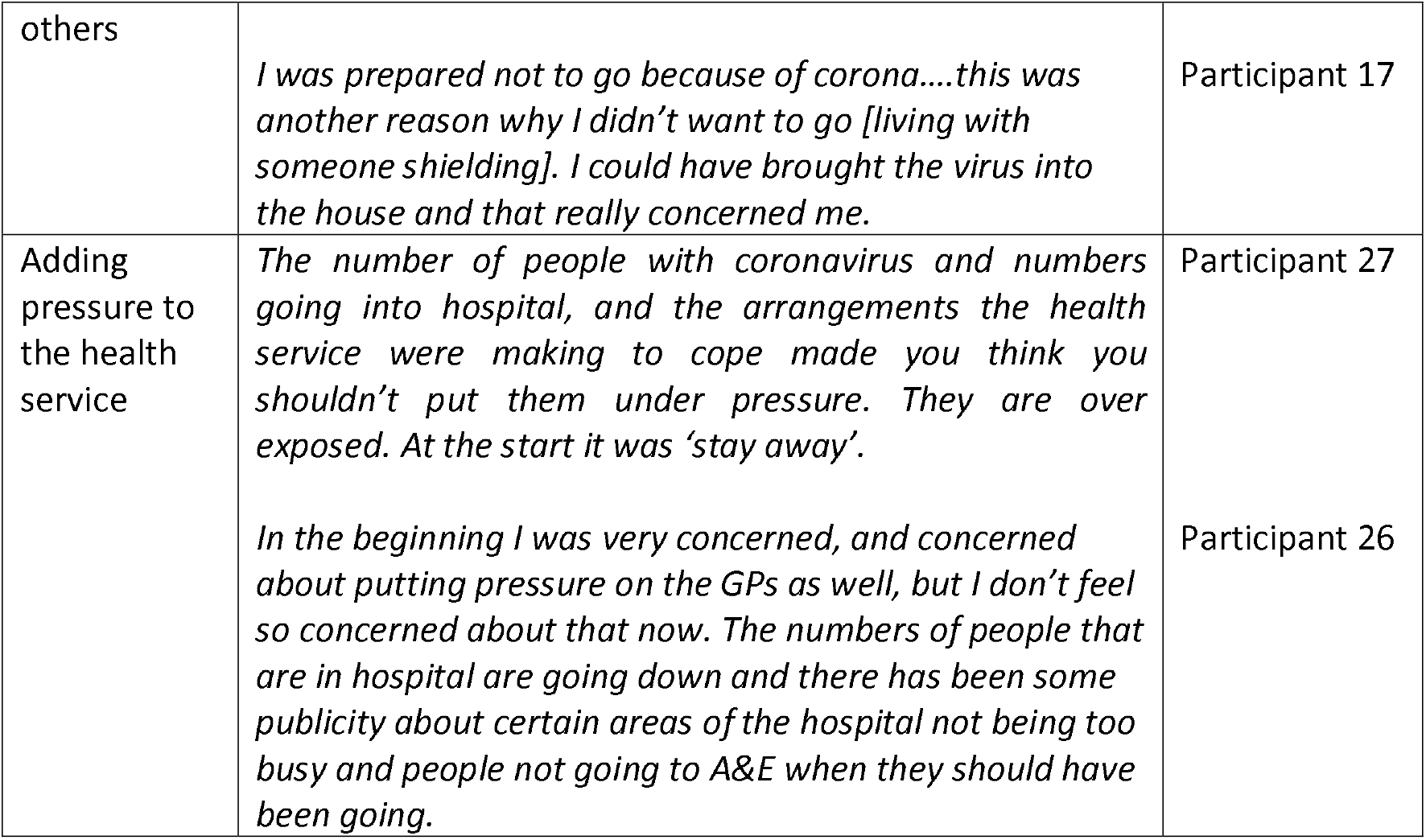
Main theme ‘COVID-19 exposure concern’.

Some participants were very concerned about presentation to hospital due to COVID-19, however, it did not stop them attending. Concerns could largely be attributed to perceived vulnerability to the virus and adding pressure to busy health services. These will be considered separately.

### Perceived vulnerability to the virus

Access to treatment and perceived vulnerability to the virus were influenced by increasing age. Some participants believed attending hospital would increase their exposure to coronavirus and discussed the possible repercussions of this with reduced access to ventilators. One participant felt at increased risk of contracting COVID-19 after sharing a hospital room with three elderly patients. He cited media reports that elderly people were more at risk of severe COVID-19 disease and interpreted that to mean a greater risk of contracting COVID-19 by sharing a room with elderly people.

Expressions of vulnerability to the virus were also seen in patients with pre-existing conditions. One participant felt vulnerable to COVID-19 due to an impaired immune system and a previous myocardial infarction. A family decision was made to limit potential exposure to coronavirus from her daughter, a nurse, by leaving the family home to live in temporary accommodation. Whilst this participant did attend hospital for assessment, she stated that she was frightened. Another previously well patient who attended hospital with myocardial infarction, stated that this acute cardiac event made him more vulnerable to coronavirus.

A further participant had two ED chest pain presentations during lockdown. Initially she was assessed and discharged then represented two weeks later with acute myocardial infarction. She knew attending was the most appropriate course of action but described an internal conversation aiming to weigh up the risks of exposing herself to the virus versus the risk of not seeking assessment for chest pain.

### Protecting others from infection

Protecting others was another consideration for participants. One participant actively wanted to attend hospital to access a test for COVID-19 so she knew she was not putting her carers at risk. Other examples included considering the exposure risk to grandchildren in the home, and inadvertently transmitting the virus from the hospital environment to the home of a vulnerable shielding adult if choosing to attend hospital.

### Adding pressure to the health service

Some participants explicitly stated they were not concerned about adding pressure to the health service. They described feeling so unwell they knew they had to attend hospital. Others had learnt that Emergency Departments were quiet through discussions with their GP or local networks which included health professionals. Participants stated they knew the hospitals were fully open.

For others, media images such as the unfolding crisis in Italy were a factor in their reluctance to attend hospital. Daily news reports detailing the number of new cases and deaths, the building of new hospitals with capacity for 5,000 patients, and images of staff wearing protective suits all contributed to the message of ‘Stay Away’ at the beginning of the pandemic. Participants stated how their perception of this message had changed over time. Publicity about decreased hospital demand was cited as a reason why some participants who would have been reluctant to use services in the beginning were now less concerned about doing so.

### Hospital experience

Participants reported a much more positive hospital experience from what they had anticipated. They stated the assessment areas were quiet, so they were seen quickly. Some were not aware that patients with COVID-19 symptoms entered the hospital and were assessed through a different point of access. Once in hospital they could see they were separated from suspected COVID-19 patients. Some participants were informed by their GP that this would be the case, others said they assumed the NHS would take this action.

*“Common sense tells you that the NHS wouldn’t be what it is unless they did keep people separate*.*” (participant 23)*

Many participants reported feeling safe while in hospital due to regular changing of personal protective equipment and hand washing by staff, in addition to highly visible cleaning taking place.

*“I felt like I was in the safest place in Edinburgh” (participant 2)*

*“It felt safer than usual. Hospitals aren’t normally the cleanest places, in my opinion, so it felt better than usual. The cleanliness and what the staff were doing was absolutely spot on. You could tell people were taking extra care” (participant 8)*

One participant commented that nurses in the hospital ward were not social distancing. It was also mentioned that not being able to have anyone accompany you to the Emergency Department or visit you in hospital made an already worrying time even more difficult.

## Discussion

Key public health strategies targeted at decreasing community transmission of SARS-CoV-2 included hand washing, social distancing and self-isolation. Mass media campaigns have previously been shown to elicit potentially beneficial behaviour change in response to the SARS and H1N1 epidemics regarding hand washing and social distancing [9]. Governments across the globe used daily briefings to keep the public informed of national developments in the fight against SARS-CoV-2 and to promote these key public health interventions. In Scotland, a slogan was developed to reiterate the fundamental message of lockdown; ‘Stay at home to protect our NHS and save lives’. Patterns of Google searches seeking advice on symptoms of chest pain and the observed reduction in acute chest pain presentations to hospital appear to align closely with the dates of public health messaging campaigns during the COVID-19 pandemic in Scotland. These findings suggest that government messaging delivered through media campaigns may have had an effect on help seeking behaviour for acute chest pain during the early stages of the COVID-19 pandemic resulting in a decrease in attendance at the Emergency Department.

The majority of participants first sought an assessment of chest pain through primary care services. Reluctance to use the emergency services has been seen prior to the COVID-19 pandemic due to concern about appropriate use of the NHS and resources [10, 11]. It appears likely that that these concerns have been enhanced during the COVID-19 pandemic. Additionally, our findings have highlighted the importance of local neighbourhood networks in providing healthcare advice out with a formal NHS framework. These networks which consisted of family, friends and neighbours working as doctors, nurses, and an acute care physiotherapist, were aware of how hospitals were managing admissions through the emergency department and were aware that local services were not nearing their maximum capacity as media reports were demonstrating both in the UK and in other countries around the world. Decision making on whether to attend hospital was therefore typically shifted to an informal health professional network, with current knowledge and understanding of how the healthcare system was functioning during the pandemic. After such informal consultations, participants were often encouraged to attend the Emergency Department for assessment. Help-seeking is described as involving three distinct elements; the person who is looking for help, the problem for which help is sought and the person from who help is required [12]. Community and social networks have previously been shown to influence help seeking [13]. Interaction with a third party may be facilitated if already known to the person requiring help. Seminal work on help seeking and social networks revealed that users of the health service prioritised daily contact with friendship groups over family groups which was reversed in non-users of the health service [14]. Social mobility has potentially enabled interactions and friendships with health professionals within local neighbourhoods. It is possible that those who chose to attend the Emergency Department during these unprecedented times were those who were able to access healthcare advice through their own informal networks. Patients with chest pain not attending for assessment may not have had the benefit of an informal local advice structure. Whether the concept of informal local healthcare networks could be exploited further to reach others during future similar healthcare crises such as the COVID-19 pandemic is an avenue for further exploration.

As the actual hospital experience was often very different to the imagined experience and largely positive, it may be useful for future media and government message campaigns to outline clearly the step by step mechanisms for people to access emergency services and to clearly describe the safety measures that Emergency Departments have taken to minimise risk for patients that need to attend hospital urgently during a future pandemic. Commercial sectors of society, for example supermarkets, have done this with television campaigns. A ‘Ways we are keeping you safe’ campaign highlighting that hospitals have taken steps to separate emergency assessment areas into COVID/non-COVID zones may make patients feel more comfortable attending the hospital.

Patient concern regarding ‘vulnerability to the virus’ has emerged as an important discourse during the pandemic due a lack of clarity on categories of patients that were and were not included in government-defined vulnerable groups. Participants frequently used their own interpretation of government messages and media articles to categorise themselves as vulnerable. Some participants were therefore faced with having to weigh up the consequences of not seeking assessment for chest pain with potential exposure to SARS-CoV-2. While communication strategies and policies are selected based on the potential for positive effect e.g. the stay at home message aiming to decrease the chance of community transmission and therefore protect vulnerable groups [15] these same strategies can produce unintended negative effects [16]. Uncertainty regarding personal vulnerability to the virus may have been exacerbated by initial uncertainty as to who was clinically vulnerable due to the unknown nature of the virus [17].

### Limitations

We were unable to access the experience of people who chose not to attend hospital with symptoms of chest pain, however, this service evaluation project has revealed valuable insights into how the decision to attend hospital was shaped by the pandemic. The first interviews were carried out at or just after the time of the release of a public health messaging campaign to promote attendance to the Emergency Department for urgent conditions. While some interviews included participants who had experienced symptoms two weeks earlier, perceived decision making may have changed in response to this new campaign. We did not explore factors influencing help seeking behaviour and hospital attendance during the early messages advising patients to stay at home and protect the NHS.

## Conclusion

We observed changes in help seeking behaviour and a reduction in attendance to hospital for the assessment of chest pain during the COVID-19 pandemic and associated lockdown in Scotland. Chest pain presentations then returned towards expected levels following public health advice to attend hospital for ‘urgent’ care. Future media and public health campaigns associated with potential second waves of COVID-19 infection should seek to strike a balance between appropriate care-seeking and avoidance behaviour. Such campaigns should be designed to include dynamic monitoring of the public response to healthcare messaging in a way that permits rapid adjustment to ensure that the net impact on health, pandemic and non-pandemic related, is optimised.

## Supporting information

Appendix 1 Topic Guide

## Data Availability

A de-identified data set can be made available for sharing upon request.

## Contributors

AVF, MAD, NLM and FS designed the project. AF and FS carried out the initial acquisition, analysis, and interpretation of data. All authors were involved in drafting and revising the manuscript and have given final approval of the version to be published. AVF takes responsibility for the paper as a whole.

