## Appendix 1 Topic Guide for "Chest pain presentations to hospital during the COVID-19 lockdown: lessons for public health media campaigns"

**Running title:** *Chest pain presentation and COVID-19*

**Address for correspondence:**

Dr Amy Ferry

BHF/University Centre for Cardiovascular Science

The University of Edinburgh

Edinburgh EH16 4SA

United Kingdom

**Appendix 1**

**Topic guide**

**Introduction**

Explain purpose of the project and ask whether the patient is happy to be involved.
State participation is voluntary.

Ask whether participant is happy for the conversation to be recorded

State the conversation will be centred around their episode of chest pain and how they made the decision to come to hospital for assessment.

(Note to interviewer – allow conversation to happen naturally. If the following information does not arise then prompts can be used.

Describe what had been happening.

Had you tried to discuss your symptoms with anyone else before attending hospital?

Were you able to access help from GP etc?

How long had you had symptoms before seeking assessment?

Did coronavirus affect your decision to attend hospital with your chest pain?

Did you know the hospital was fully open to assess patients?

Were you concerned about coming into hospital due to coronavirus?

What were you concerned about 1) overwhelming the health service 2) being exposed to coronavirus?

Was your concern for yourself or someone you live with?

What made you think you should not attend hospital? (are official media messages mentioned?)

What was your experience of being in hospital for chest pain assessment? Did you feel safe?

Was is different to what you had expected?

If we could offer a video call assessment for your symptoms would that service be useful?

If you had a further episode of chest pain what would your action be?
